# Orthopoxvirus-specific antibodies wane to undetectable levels one year after MVA-BN vaccination of at-risk individuals

**DOI:** 10.1101/2024.08.13.24311601

**Authors:** Leanne P. M. van Leeuwen, Marc C. Shamier, Babs E. Verstrepen, Hannelore M. Götz, Katharina S. Schmitz, Najlae Akhiyate, Koen Wijnans, Susanne Bogers, Martin E. van Royen, Eric C. M. van Gorp, Marion P. G. Koopmans, Rory D. de Vries, Corine H. GeurtsvanKessel, Luca M. Zaeck

**Affiliations:** Department of Viroscience, Erasmus University Medical Center, Rotterdam, the Netherlands; Travel Clinic, Erasmus University Medical Center Rotterdam, the Netherlands; Department Infectious Disease Control, Municipal Public Health Service Rotterdam–Rijnmond (GGD Rotterdam-Rijnmond), Rotterdam, the Netherlands; Department of Pathology, Erasmus University Medical Center, Rotterdam, the Netherlands

## Abstract

In response to the 2022-2023 mpox outbreak, widespread vaccination with modified vaccinia Ankara-Bavarian Nordic was initiated. Here, we demonstrate that orthopoxvirus-specific binding and MVA-neutralizing antibodies wane to undetectable levels one year post-vaccination in at-risk individuals without pre-existing immunity. Continuous surveillance is essential to understand the impact of declining antibody levels.

## Introduction

Since the beginning of the 2022-2023 global mpox outbreak, over 97,000 cases were reported in 117 countries, including in previously non-endemic regions [1]. The vast majority of monkeypox virus (MPXV) infections outside endemic regions were diagnosed in sexually active gay, bisexual, or other men who have sex with men (GBMSM). To control the outbreak, many countries initiated vaccination campaigns using the third-generation smallpox vaccine modified vaccinia Ankara – Bavarian Nordic (MVA-BN). In the initial months after the start of vaccination, studies on vaccine effectiveness (VE) yielded an estimated aggregate VE of 82% (CI: 65%-89%) after two doses [2]. Breakthrough infections after vaccination have been reported [3]. Combined, this raises questions on the long-term effectiveness of MVA-BN against mpox.

We have previously shown that a two-dose MVA-BN vaccination regimen is immunogenic in individuals without pre-existing immunity; vaccination resulted in the production of VACV (vaccinia virus)-reactive binding antibodies, MVA-neutralizing antibodies (nAbs), and MPXV nAbs one month after the second dose. However, the levels of MPXV nAbs were low compared to those after historic smallpox vaccination, or after MPXV infection [4]. Little is known about the durability of humoral immune responses, and the correlation between waning antibody responses and long-term effectiveness of MVA-BN vaccination. While antibodies induced by historic smallpox vaccination using first- or second-generation vaccines can be detected for decades [5], prior studies into the longevity of orthopoxvirus-specific antibodies after MVA-BN vaccination indicate that antibodies wane in vaccine recipients without pre-existing immunity [6-10].

In this study, we specifically investigated the durability of orthopoxvirus-specific antibody responses and report the longitudinal antibody dynamics up to 418 days after MVA-BN vaccination in two risk groups, namely GBMSM and laboratory workers.

## Methods

Sera from two biobank protocols were used. Both protocols were approved by the Erasmus MC Medical Ethics Review Committee: the COVA study (MEC-2014-398) and the RestPlus study (MEC-2022-0675) (**Supplementary Methods** and **Supplementary Figure S1**). Written informed consent was obtained from all COVA study participants. Baseline characteristics are documented in **Supplementary Table S1**.

In the COVA study, sera were collected from both high-risk GBMSM (N=99) and laboratory workers (N=19). Laboratory workers were vaccinated as part of occupational safety measures. Samples were obtained before and after MVA-BN vaccination (V0=baseline; V1=14 days after first dose; V2=28 days after first dose; V3=28 days after second dose; V4=1 year follow-up) (**Supplementary Table S2A)**. Participants were vaccinated according to the prescribed vaccination regimen: two subcutaneous doses with an interval of four weeks for individuals who had not received historic smallpox vaccination. This included participants born after 1974 (cessation of smallpox vaccination in the Netherlands), and those born before 1974 with unclear vaccination history. Laboratory workers received two doses of MVA-BN regardless of vaccination history. Other individuals who declared having received historic smallpox vaccination received one MVA-BN dose. In the RestPlus study, residual pseudonymized sera obtained in September 2022 from GBMSM attending the Center for Sexual Health (CSH) in Rotterdam were screened for the presence of VACV-specific binding antibodies [11]. In case of a positive result, all sera from that individual collected between January 2022 and September 2023 were tested, and data were linked to available vaccination- and/or infection data. Individuals without registration of an administered MVA-BN vaccine, or without a positive MPXV PCR, were excluded from further analysis. Longitudinal sera from N=116 individuals were eligible for analysis.

We measured VACV-specific binding antibody levels using an in-house ELISA in both cohorts. For sera obtained from the COVA cohort, MVA nAb levels were measured by plaque reduction neutralization test (PRNT). Both assays were performed as previously described [4] (**Supplementary Methods**).

## Results

In total, we analyzed sera from 234 participants enrolled in two different studies. Of the 99 GBMSM and 19 laboratory workers enrolled in the COVA study, 17 participants received one dose and 101 participants received two doses of MVA-BN (**Supplementary Figure S1** and **Supplementary Table S1**). As expected, no VACV-specific binding antibodies were detected at baseline in either GBMSM or laboratory workers born after 1974 (**Figure 1A**). Binding antibody levels gradually increased over time with detectable antibodies in all sera (58/58; 100%) collected four weeks post-2^nd^ dose (GMT 160 [95% CI 116-220]). However, only 7/21 (33%) participants had detectable VACV-specific binding antibody levels above the lower limit of detection (LLoD) at one year post-2^nd^ dose (GMT 15 [95% CI 11-21]). We did not observe a difference in binding antibody kinetics post-vaccination between GBMSM and laboratory workers (**Supplementary Figure S2)**.

**Figure 1.**
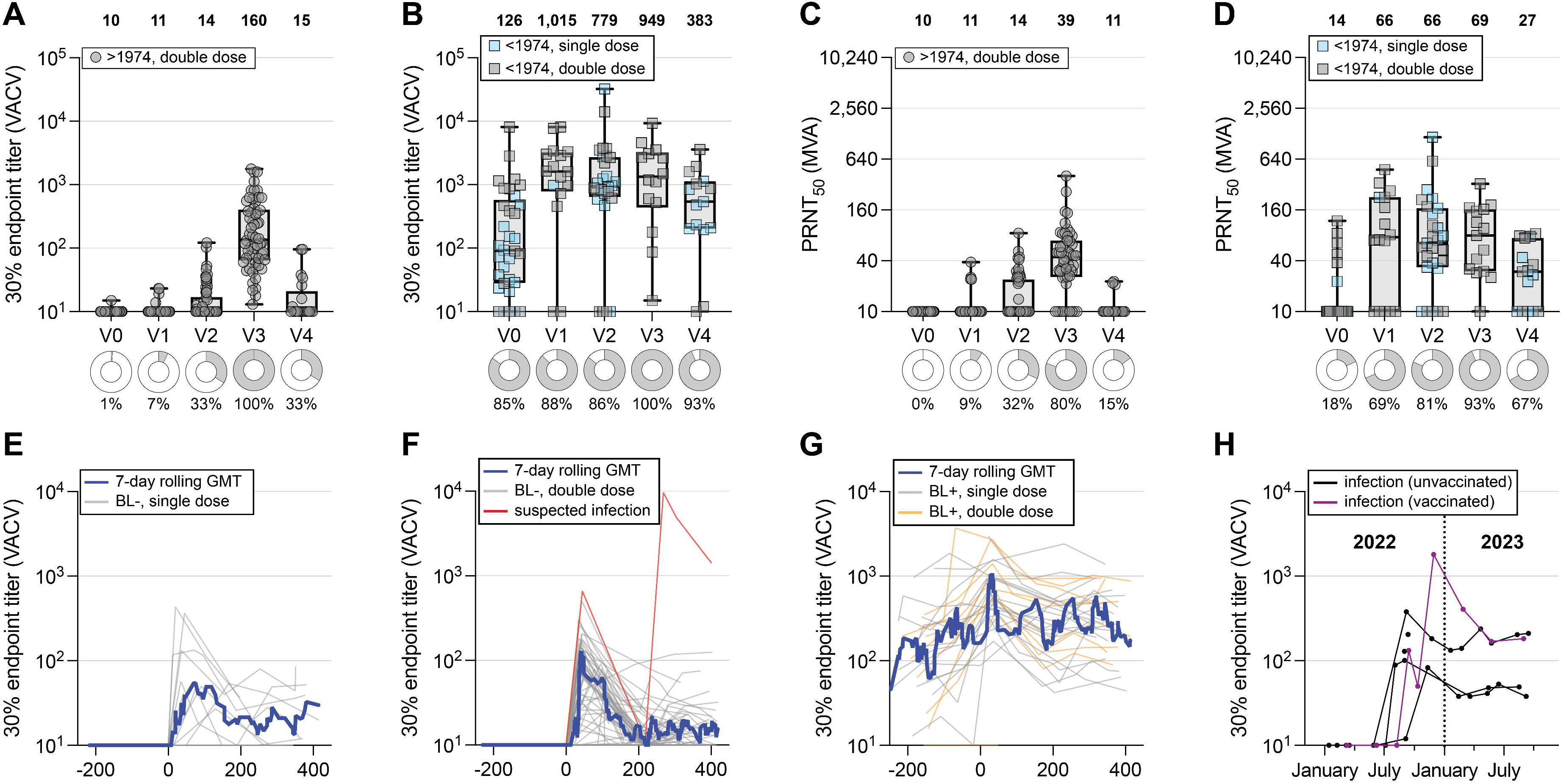
Antibody longevity after MVA-BN vaccination. VACV-specific binding or MVA neutralizing antibodies (nAbs) were measured in sera obtained from two cohorts: **(A-D)** COVA (individuals born from 1974 onwards are presumed to be immunologically naive, those born before 1974 are presumed to have received historic smallpox vaccination); and **(E-H)** RestPlus. **(A)** VACV-specific binding antibody levels in immunologically naive participants (circles) after receiving two doses of MVA-BN. **(B)** VACV-specific binding antibody levels in participants with pre-existing immunity (squares) after receiving either one dose (blue) or two doses (gray) of MVA-BN. One participant born before 1974 deviated from the study protocol and had a V1 instead of a V2 visit. **(C, D)** MVA nAbs in naive participants **(C)** or participants with pre-existing immunity **(D)**, after receiving one (blue) or two doses (gray) of MVA-BN. Data are shown in box-and-whisker plots, with the horizontal lines indicating the median, the bounds of the boxes indicating the interquartile range (IQR), and the whiskers indicating the range. Bold numbers above the plots represent the geometric mean titer (GMT) per timepoint. Donut graphs show percentage of positive samples per timepoint. **(E, F)** VACV-specific binding antibody levels in RestPlus participants without detectable antibodies at baseline (BL-) after receiving either one (**E**) or two doses of MVA-BN **(F)**. The red line represents a suspected infection. **(G)** VACV-specific binding antibody levels in RestPlus participants with detectable antibodies at baseline (BL+) and either one (gray line) or two doses (orange line) of MVA-BN. **(H)** VACV-specific binding antibody levels in RestPlus participants with a PCR-confirmed MPXV infection. All individuals were unvaccinated except for one (purple line). The blue line in **E-G** represents the 7-day rolling GMT; V0=baseline; V1=14 days after first dose; V2=28 days after first dose; V3=28 days after second dose; V4=1 year follow-up.

In individuals born before 1974, VACV-specific binding antibodies were detected at baseline before MVA-BN vaccination in 28/33 participants (85%) (**Figure 1B** and **Supplementary Table S2B**). Binding antibody levels were boosted by the first dose of MVA-BN (GMT 897 [95% CI 347-2,324] for those receiving one dose, and GMT 675 [95% CI 166-2,742] for those receiving two doses) at 28 days after first vaccination. A second dose of MVA-BN did not result in an additional increase of binding antibody levels (GMT 949 [95% CI 347-2,596] at 28 days after second vaccination; **Figure 1B**). Binding antibody levels in MVA-BN-vaccinated participants with prior immunity remained stable at the one-year follow-up. Two participants consistently had low or undetectable antibody levels.

To assess the durability of antibody functionality, we measured MVA nAbs in participants of the COVA study. In participants born after 1974, we detected MVA nAbs in 41/51 (80%) study participants 28 days after the second MVA-BN vaccination (GMT 39 [95% CI 31-51]), compared to only 3/20 (15%) participants one year post-vaccination (GMT 11 [95% CI 9.8-13]) (**Figure 1C**). In participants born before 1974, MVA nAbs were boosted by the first dose of MVA-BN, without an additional increase after the second dose, comparable to binding antibodies (**Figure 1D**). One year post-vaccination, MVA nAb levels waned, with 5/15 (33%) individuals having MVA nAb levels below the LLoD.

Antibody kinetics were additionally studied up to 418 days post-vaccination in longitudinal serum samples obtained from 116 GBMSM clients of the Rotterdam CSH, who received MVA-BN vaccination (N=100) or had a positive MPXV PCR (N=6) (RestPlus study) (**Supplementary Table S2C**). The vaccinated participants were grouped based on the absence or presence of VACV-specific binding antibodies on the day of MVA-BN vaccination into a baseline-negative (**Figure 1E** [one dose]; **Figure 1F** [two doses]) and a baseline-positive (**Figure 1G**) group. Independent of the status at baseline, all groups showed an increase in binding antibody levels post-vaccination, which peaked within 30-90 days. Subsequently, binding antibody levels began to decline across all groups, returning close to their respective baseline levels one year post-vaccination. For baseline-negative study participants, among those who were followed-up for a year or longer, 65% (22/34) had low (<20) to no detectable antibodies at the end of the study, whereas baseline-positive individuals returned to their original baseline levels. Overall, this longitudinal cohort mirrored antibody kinetics observed in the COVA study.

In the cohort of GBMSM clients of the Rotterdam CSH, there were six participants with a PCR-confirmed mpox during the study period, all born after 1974 (**Figure 1H**). For two of these individuals, only a single serum sample was available. One participant was diagnosed with an MPXV infection in October 2022, following the completion of a double-dose MVA-BN vaccination regimen in early September 2022 (**Figure 1H**, purple line). Comparison of binding antibody levels shortly before and one month after infection revealed an increase from 50 to 1807. Based on the binding antibody kinetics and magnitude, we suspect an additional undetected infection among one of the participants after two doses of MVA-BN (**Figure 1G**, red line). However, no clinical information and/or confirmatory PCR-tests were available for this participant.

## Discussion

Here, we showed that MVA-BN vaccination induced binding and MVA neutralizing antibodies in previously unvaccinated at-risk study participants (born ≥1974), and that vaccination boosts binding antibody levels in historically vaccinated at-risk individuals (born <1974). Binding and neutralizing antibody levels in non-primed at-risk individuals declined rapidly at one year follow-up, and became undetectable in a considerable proportion of cases.

Measuring the immunogenicity of MVA-BN is crucial for understanding the impact of immunization strategies and supporting VE studies. Previous reports on long-term humoral immunogenicity align with our findings [6-10]. However, we expand on them by providing longitudinal immunological follow-up of over one year in those directly at-risk of clade IIb MPXV infection. While the implications of low to absent antibody levels one-year post vaccination remain unclear due to the absence of a clearly defined correlate of protection, waning antibody levels raise the important question whether these individuals are still protected and if this could possibly facilitate mpox resurgence.

In our study, we observed one confirmed and one suspected breakthrough infection, both associated with an increase in binding antibody levels. Breakthrough infections after MVA-BN vaccination have been previously reported and appear to result in a milder clinical profile [3]. Explanations for breakthrough infections shortly after vaccination could be due to antigenic differences between MVA and MPXV that result in suboptimal cross-reactivity, or the potential absence of immunity at mucosal sites, a point of entry for clade IIb MPXV [4-10].

We demonstrate a rapid decline in binding and neutralizing antibodies one-year after MVA-BN vaccination in non-primed at-risk individuals. Combined with the continuing burden of MPXV, this highlights the importance of further investigating the long-term efficacy of MVA-BN. Ongoing circulation of clade I(b) MPXV in the Democratic Republic of the Congo [12] necessitates future studies elaborating on the cross-reactivity of vaccine-induced antibody responses against this circulating clade. In addition, administration of a booster dose one year after the initial dose could be considered to improve vaccine immunogenicity [4, 6]. Taken together, our data contribute to the understanding of waning humoral immune responses following MVA-BN vaccination, and support decision making on vaccination strategies.

## Data Availability

All data produced in the present study are available upon reasonable request to the authors.

## Acknowledgments

The authors thank the colleagues of the GGD Rotterdam-Rijnmond (Municipal Public Health Service Rotterdam-Rijnmond, Rotterdam, the Netherlands) and the Travel Clinic Erasmus MC who supported this study. We thank Denise Twisk (Municipal Public Health Service Rotterdam-Rijnmond, Rotterdam, the Netherlands) for help with management of clinical data. This project has received funding from the European Union’s funding program H2020 research and innovation program under grant agreement 101115188 and the Netherlands Organization for Health Research and Development (ZonMw) under grant agreement 10150022310035.

## Conflicts of interest

No reported conflicts of interest.

## Author contributions

Study conceptualization: LPMvL, MCS, BEV, HMG, ECMG, RDdV, CHGvK, LMZ. Methodology: LPMvL, MCS, BEV, ECMG, RDdV, CHGvK, LMZ. Formal analysis: LPMvL, MCS, BEV, RDdV, CHGvK, LMZ. Investigation: LPMvL, MCS, BEV, HMG, NA, KSS, KW, SB, ECMG, RDdV, CHGvK, LMZ. Resources: HMG, MPGK, ECMvG, RDdV, CHGvK. Data curation: LPMvL, MCS, BEV, ECMG, RDdV, CHGvK, LMZ. Project administration: LPMvL, MCS, BEV, ECMG, RDdV, CHGvK, LMZ. Visualization: LPMvL, MCS, LMZ. Supervision: ECMvG, RDdV, CHGvK, LMZ. Writing – original draft: LPMvL, MCS, BEV, RDdV, CHGvK, LMZ. Writing – review and editing: All authors.

## Notes

### Competing Interest Statement

The authors have declared no competing interest.

### Author Declarations

Sera from two biobank protocols were used. Both protocols were approved by the Erasmus MC Medical Ethics Review Committee: the COVA study (MEC-2014-398) and the RestPlus study (MEC-2022-0675).

## References

1. WHO. Multi-country outbreak of mpox. External Situation Report 33. Available at: https://www.who.int/publications/m/item/multi-country-outbreak-of-mpox--external-situation-report-33--31-may-2024. Accessed 24-06-2024.

2. Berry MT, Khan SR, Schlub TE, et al. Predicting vaccine effectiveness for mpox. Nat Commun 2024; 15(1): 3856.

3. Hazra A, Zucker J, Bell E, et al. Mpox in people with past infection or a complete vaccination course: a global case series. Lancet Infect Dis 2024; 24(1): 57–64.

4. Zaeck LM, Lamers MM, Verstrepen BE, et al. Low levels of monkeypox virus neutralizing antibodies after MVA-BN vaccination in healthy individuals. Nat Med 2022.

5. Taub DD, Ershler WB, Janowski M, et al. Immunity from smallpox vaccine persists for decades: a longitudinal study. Am J Med 2008; 121(12): 1058–64.

6. Ilchmann H, Samy N, Reichhardt D, et al. One- and Two-Dose Vaccinations With Modified Vaccinia Ankara-Bavarian Nordic Induce Durable B-Cell Memory Responses Comparable to Replicating Smallpox Vaccines. J Infect Dis 2023; 227(10): 1203–13.

7. Oom AL, Kottkamp AC, Wilson KK, et al. The Durability and Avidity of MPXV-specific Antibodies Induced by the Two-dose MVA-BN Mpox Vaccine. medRxiv 2024: 2024.01.28.24301893.

8. Moraes-Cardoso I, Benet S, Carabelli J, et al. Immune responses associated with mpox viral clearance in men with and without HIV in Spain: a multisite, observational, prospective cohort study. Lancet Microbe 2024.

9. Priyamvada L, Carson WC, Ortega E, et al. Serological responses to the MVA-based JYNNEOS monkeypox vaccine in a cohort of participants from the Democratic Republic of Congo. Vaccine 2022; 40(50): 7321–7.

10. Crandell J, Monteiro VS, Pischel L, et al. The impact of antigenic distance on Orthopoxvirus Vaccination and Mpox Infection for cross-protective immunity. medRxiv 2024: 2024.01.31.24302065.

11. Shamier MC, Zaeck LM, Götz HM, et al. Scenarios of future mpox outbreaks among men who have sex with men: a modelling study based on cross-sectional seroprevalence data from the Netherlands, 2022. Euro Surveill 2024; 29(17).

12. Vakaniaki EH, Kacita C, Kinganda-Lusamaki E, et al. Sustained human outbreak of a new MPXV clade I lineage in eastern Democratic Republic of the Congo. Nat Med 2024.

