## Supplementary Material for "Orthopoxvirus-specific antibodies wane to undetectable levels one year after MVA-BN vaccination of at-risk individuals"

* Equal first author contribution

[#] Equal senior author contribution

### Supplementary Methods

#### Study design

The sera used in this study were derived from two different studies, namely COVA and RestPlus.

COVA

Individuals who received the modified vaccinia Ankara – Bavarian Nordic (MVA-BN) vaccine were selected from the vaccination cohort biobank. From August 2022 onwards, 118 individuals who received the MVA-BN vaccine had given written informed consent to participate in the biobank study. These individuals belonged to two different risk groups: sexually active gay, bisexual, or other men who have sex with men (GBMSM) (N = 99) and laboratory workers (N=19) (**Supplementary Figure S1** and **Supplementary Table S1**). No study participant reported a previous infection with mpox at the time of inclusion.

According to Dutch guidelines, pre-exposure MVA-BN vaccination was offered to high-risk GBMSM who were selected based on a recent sexually transmitted infection (STI) or who met the criteria to use HIV pre-exposure prophylaxis (PrEP). The MVA-BN vaccination regimen consisted of two vaccinations with an interval of four weeks for individuals who had not received historic smallpox vaccination. As routine smallpox vaccination in the Netherlands was stopped in 1974, this included participants born after 1974 and those born before 1974 with unclear vaccination records. Individuals born before 1974 who had received historic childhood smallpox vaccination received one MVA-BN dose. Laboratory workers received two doses of MVA-BN regardless of prior vaccination history. All doses were given subcutaneously and contained a minimal amount of $5 \times 10^7$ plaque-forming units. According to the protocol of the COVA study, a maximum of five blood samples could be collected before and after the administration of a non-experimental vaccine. Longitudinal blood samples of participants receiving two vaccines were collected at baseline (V0), 14 days after the first vaccination (V1), 28 days after the first vaccination (before the administration of the second vaccine; V2), 28 days after the second vaccination (V3), and one year after second vaccination (V4). Three blood samples were collected from individuals who received a single dose of the MVA-BN vaccine: at baseline (V0), 28 days after vaccination (V2), and one year after vaccination (V4). Blood samples from time points V0-V3 of the laboratory workers were collected using EDTA tubes; all other time points and all blood samples from GBMSM were collected using serum tubes.

Clinical data were obtained via questionnaires, which were stored in a secure electronic data management platform (CASTOR EDC). Clinical data and samples were stored pseudonymized. Due to the biobank format of the COVA study, it has been exempted from medical ethical approval. This exemption has been confirmed by the Erasmus MC Medical Ethics Review Committee (MEC-2014-398).

RestPlus

Residual serum samples obtained from GBMSM at the Center for Sexual Health (CSH) of the Rotterdam Municipal Health Department underwent testing for vaccinia virus (VACV)-specific binding antibodies at the Department of Viroscience of the Erasmus University Medical Center, Rotterdam.

Initially, all residual samples collected in September 2022 were screened as part of a serosurveillance study [1]. In case of a positive result for VACV-specific antibodies, subsequent testing extended to all available residual sera collected from January 2022 to December 2023 from the same individual (**Supplementary Figure S1** and **Supplementary Table S1**). In case of undetectable VACV-specific antibodies, samples were excluded from further analysis. In total, 143 individuals of 315 individuals tested positive for the presence of VACV-specific antibodies. After a privacy impact assessment was conducted and a data sharing agreement was signed between the Center for Sexual Health (CSH) of the Rotterdam Municipal Health Department and the Department of Viroscience of the Erasmus University Medical Center, serology data were combined with MVA-BN vaccination data and mpox infection status. Individuals without registration of either the administration of MVA-BN or a PCR-confirmed monkeypox virus (MPXV) infection were excluded from further analysis (N=27).

The longitudinal sets of residual sera of the remaining 116 individuals (N=110 with records of administered MVA-BN vaccination(s) and N=6 with PCR-confirmed MPXV infection) were tested under the study protocol RestPlus (MEC-2022-0675). This protocol was reviewed and approved by the Erasmus MC Medical Ethics Review Committee. Donors have not been asked to donate additional samples for the purpose of this study at any time point. Only the donors' year of birth, gender, positive MPXV PCR tests, and MVA-BN vaccination records were available. These data were stored pseudonymized.

**Detection of vaccinia virus (VACV)-specific IgG antibodies by ELISA**

Binding IgG antibody levels against VACV were measured using an in-house developed ELISA as described previously [2]. Briefly, high-binding 96-well ELISA plates (Corning) were coated with the cell lysate of either VACV-infected or mock-treated HeLa cells diluted 1:250 in PBS for 1 h at 37 °C. After coating, plates were washed with washing buffer (PBS + 0.05% Tween20 [Merck]) and blocked with for 1 h at 37 °C using blocking buffer (2% skim milk powder [Merck] in washing buffer). Fivefold dilution series of the sera in blocking buffer starting at a 1:10 dilution were prepared, added to the plates, and incubated overnight at 4 °C. Afterwards, plates were washed with washing buffer and incubated for 1 h at 37 °C with horseradish peroxidase (HRP)-conjugated rabbit anti-human IgG (1:6,000; Dako) diluted in blocking buffer. After another washing step, plates were developed with TMB peroxidase substrate (SeraCare). Absorbance was measured at 450 nm using a microtiter plate reader (Anthos 2001 microplate reader) and corrected by subtracting absorbance at 620 nm. A net $OD_{450}$ response was determined by subtracting the $OD_{450}$ value from plates coated with mock-treated cell lysate from those coated with VACV-infected cell lysate. A positive reference serum pool was included on every plate and used to calculate 30% endpoint titers by transforming the net $OD_{450}$ responses per sample to this positive control S-curve. Short-term results of the laboratory worker cohort (up to V3; n = 14) were published previously and are displayed again to reflect the kinetics of the antibodies one year after MVA-BN vaccination [2].

**Detection of modified vaccinia Ankara (MVA) neutralizing antibodies by PRNT**

A plaque reduction neutralization test (PRNT) was used to test serum samples for their capacity to neutralize MVA as previously described [2]. Briefly, Vero cells were seeded 1 day before the experiment in 96-well cell culture plates. Heat-inactivated sera were twofold serially diluted in Advanced DMEM/F-12 (Gibco) supplemented with 10 mM HEPES, 1× GlutaMAX and 1× penicillin/streptomycin before adding an equal volume of medium containing 1,500 PFU rMVA-GFP. The virus-serum mix was incubated for 1 h at 37 °C before being transferred to the cell culture plates. After incubation for 24 h at 37 °C and 5% $CO_2$, the cells were fixed with 4% paraformaldehyde for 10 min. The cells were washed with PBS and cell nuclei were counterstained with Hoechst33342 (Thermo Fisher Scientific). The cells were imaged using the Opera Phenix spinning disk confocal HCS system (PerkinElmer) equipped with a ×10 air objective (NA 0.3) and 405-nm and 488-nm solid-state lasers. Infected cells were quantified using the Harmony software (version 4.9, PerkinElmer). The dilution that would yield 50% reduction of plaques compared with the infection control was estimated by determining the proportionate distance between two dilutions from which an endpoint titer was calculated. If no neutralization was measured, the $PRNT_{50}$ value was assigned as 10, one dilution factor below the lowest dilution tested.

**Statistical analysis**

Baseline characteristics were described for the participants of the COVA and RestPlus study separately (**Supplementary Table S1**). Categorical variables were displayed as counts and percentages. Ages and time intervals were displayed as medians and interquartile ranges (IQR). Binding and neutralizing antibody levels were expressed as geometric mean titers (GMT) and 95% confidence intervals (95% CI). Graphs were created using GraphPad Prism (version 10.2.2).

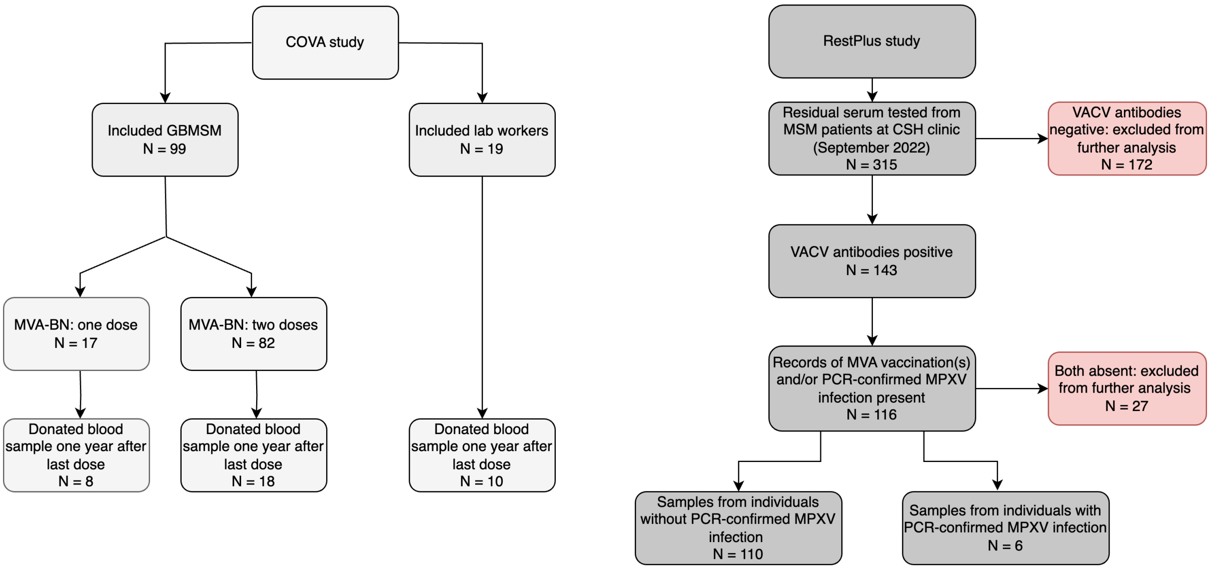

**Supplementary Figure S1. Flowcharts of the COVA study and RestPlus study**. Participants of the COVA study (left) belonged to two different risk groups: sexually active gay, bisexual, or other men who have sex with men (GBMSM) and laboratory workers. The RestPlus study (right) was performed on residual serum samples obtained from GBMSM at the Center for Sexual Health (CSH) of the Municipal Public Health Department Rotterdam-Rijnmond. Samples were eligible if they tested positive for VACV-specific antibodies and if clinical records of administered MVA vaccination(s) and/or PCR-confirmed MPXV infections were available. Six individuals included in the study had a PCR-confirmed MPXV infection. One of these six also received two MVA vaccinations before infection (indicated as purple line in **Figure 1H**).

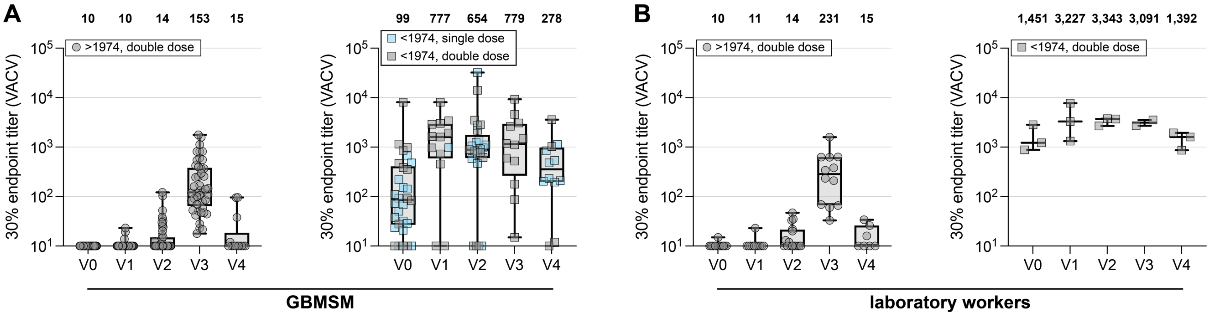

**Supplementary Figure S2. Antibody longevity after MVA-BN vaccination stratified by study cohort of the COVA study.** VACV-specific binding antibody titers were measured in sera obtained from the COVA study and stratified by study group. **(A)** VACV-specific binding antibody levels in GBMSM (gay, bisexual and other men who have sex with men) born after (left, circles) or before 1974 (right, squares) after receiving either one (blue) or two doses (gray) of MVA-BN. **(B)** VACV-specific binding antibody levels in laboratory workers born after (left, circles) or before 1974 (right, squares) after receiving two doses (gray) of MVA-BN. Administration of historic smallpox vaccination was inferred in those born before 1974 after which it was stopped in the Netherlands. Data are shown in box-and-whisker plots, with the horizontal lines indicating the median, the bounds of the boxes indicating the interquartile range (IQR), and the whiskers indicating the range. Bold numbers above the plots represent the geometric mean titer per timepoint. V0=baseline; V1=14 days after first dose; V2=28 days after first dose; V3=28 days after second dose; V4=1 year follow-up.

**Supplementary Table 1**: Baseline characteristics.

| | COVA study | | | RestPlus study | | |
|---|---|---|---|---|---|---|
| | GBMSM (N=99) | | Laboratory workers (N=19) | MVA-vaccinated or infected GBMSM (N=116) | | |
| | Single dose group N=17 (17%) | Double dose N=82 (83%) | Double dose N=19 (100%) | Sero-negative at baseline (N=78) | Sero-positive at baseline (N=32) | PCR-confirmed infection (N=6) |
| **Age (in 2022)** Median (IQR) | 56 (51-61) | 35 (28-44) | 33 (30-42) | 33 (28-40) | 56 (52-61) | 36 (35-37) |
| ≥1974: 18 – 48 yr | 0 | 69 | 16 | 78 | 2 | 6 |
| <1974: 49 – 59 yr | 10 | 9 | 2 | 0 | 18 | 0 |
| <1974: ≥ 60 yr | 7 | 4 | 1 | 0 | 12 | 0 |
| **Gender, male** | 17 (100%) | 82 (100%) | 8 (42%) | 78 (100%) | 32 (100%) | 6 (100%) |
| **HIV Status** | | | | | | |
| Positive | 2 (12%) | 3 (4%) | 0 | 0 | 0 | 0 |
| Negative | 15 (88%) | 76 (93%) | 17 (90%) | 0 | 0 | 0 |
| Unknown | 0 | 3 (4%) | 2 (10%) | 78 (100%) | 32 (100%) | 6 (100%) |
| **Medical history** | | | | NA | NA | NA |
| No comorbidity | 10 | 57 | 16 | | | |
| Hypertension | 1 | 8 | 1 | | | |
| Asthma | 1 | 2 | 1 | | | |
| Diabetes mellitus type 1 | | 3 | | | | |
| Other autoimmune diseases | 1 | 2 | 2 | | | |
| Malignity | 1 | | | | | |
| Other comorbidity* | 3 | 11 | 1 | | | |
| **PCR-confirmed mpox infection** | | | | | | |
| MVA-BN unvaccinated | 0 | 0 | 0 | 0 | 0 | 5 |
| MVA-BN vaccinated | 0 | 0 | 0 | 0 | 0 | 1 |

*Other comorbidity: ADHD (N=4), epilepsy (N=1), migraine (N=1), herpes infection (N=1), not further specified (N=8). GBMSM = gay, bisexual and other men who have sex with men; MVA = modified vaccinia Ankara.

**Supplementary Table S2. Timing and geometric mean titers of the COVA and RestPlus study.**

**Supplementary Table S2A.** Timing of study visits in the COVA study.

| | Study visit | | | | |
|---|---|---|---|---|---|
| | **V0** median (IQR) | **V1** median (IQR) | **V2** median (IQR) | **V3** median (IQR) | **V4** median (IQR) |
| **Days from V0** | N/A | 14 (14-16) | 28 (28-32) | 57 (56-63) | 393 (392-447) |

N/A = not applicable; IQR = interquartile range.

**Supplementary Table S2B.** VACV-specific 30% endpoint titers in the COVA study.

| | Study visit | | | | |
|---|---|---|---|---|---|
| | **V0** GMT (95% CI) | **V1** GMT (95% CI) | **V2** GMT (95% CI) | **V3** GMT (95% CI) | **V4** GMT (95% CI) |
| **Born ≥1974 (2 doses), N=85** | 10 (10-10) | 11 (10-11) | 14 (12-16) | 160 (116-220) | 15 (11-21) |
| **Born <1974 (1 dose), N=17** | 65 (35-121) | 973 (N/A) | 897 (347-2,324) | N/A | 385 (214-691) |
| **Born <1974 (2 doses), N=16** | 255 (83-785) | 1,018 (330-3,141) | 675 (166-2,742) | 949 (347-2,596) | 382 (39-3,751) |

N/A = not applicable; GMT = geometric mean titer; 95% CI = 95% confidence interval; V0=baseline; V1=14 days after first dose; V2=28 days after first dose; V3=28 days after second dose; V4=1 year follow-up. One participant born before 1974 deviated from the study protocol and had a V1 visit instead of a V2 visit.

**Supplementary Table S2C.** VACV-specific 30% endpoint titers in the RestPlus study.

| | Time windows normalized by date of first vaccine dose (days) | | | | |
|---|---|---|---|---|---|
| | **≤0 days** GMT (95% CI) | **1 – 90 days** GMT (95% CI) | **91 – 180 days** GMT (95% CI) | **181 – 365 days** GMT (95% CI) | **>365 days** GMT (95% CI) |
| **Seronegative at baseline (1 dose), N=19** | 10 (10-10) | 26 (14-45) | 29 (14-59) | 19 (14-26) | 26 (9.2-76) |
| **Seronegative at baseline (2 doses), N=59** | 10 (10-10) | 46 (34-62) | 22 (16-29) | 17 (14-20) | 19 (13-28) |
| **Seropositive at baseline (1 or 2 doses), N=32** | 160 (141-181) | 529 (433-646) | 303 (248-371) | 293 (268-321) | 202 (179-228) |

GMT = geometric mean titer; 95% CI = 95% confidence interval.

### References

1. Shamier MC, Zaeck LM, Götz HM, et al. Scenarios of future mpox outbreaks among men who have sex with men: a modelling study based on cross-sectional seroprevalence data from the Netherlands, 2022. Euro Surveill **2024**; 29(17).
2. Zaeck LM, Lamers MM, Verstrepen BE, et al. Low levels of monkeypox virus neutralizing antibodies after MVA-BN vaccination in healthy individuals. Nat Med **2022**.
